# Current substance use patterns and associated factors among Ghanaian adolescents in senior high school

**DOI:** 10.1101/2024.07.18.24310635

**Authors:** Rachael Asantewaa Darko, Franklin N. Glozah

**Affiliations:** Department of Social and Behavioural Sciences, School of Public Health, University of Ghana; Food and Drugs Authority, Accra, Ghana

**Keywords:** Substance use, Adolescents, Ghana, Risk factors

## Abstract

Substance use poses a significant threat to adolescent health and well-being globally, with rising rates of concern in developing countries. Understanding the specific factors currently driving substance use among youth is crucial for developing targeted interventions. This study examines current substance use patterns and their correlates among Ghanaian in-school adolescents. A cross-sectional survey was administered to a random sample of 425 SHS students in Accra. A standardised questionnaire was used to assess substance use patterns (types, frequency, age of initiation), peer and family influences, socio-demographic characteristics and potential substance dependence. Descriptive statistics were used to characterize the sample and a multiple logistic regression models identified predictors of use for specific substances. The mean age of participants was 17.1 years. Cigarettes were the most used substance, followed by shisha, marijuana, and alcohol. Male students, those living with relatives, and those with friends who drink alcohol were more likely to use alcohol. Students who worked while in school, or had family members who smoke, had an increased likelihood of cigarette use. Older students and those with friends who use shisha were more likely to use shisha. Interestingly, limited social media exposure and living with parents and siblings were associated with lower marijuana use. Additionally, the results showed a potential substance dependence in some students. This study highlights substance use patterns and influential factors among adolescents in urban Ghana. Findings emphasize the interaction of peer influence, family environment, and gender in shaping substance use behaviours. These insights can inform culturally sensitive interventions to promote adolescent health and resilience in Ghana, and potentially other developing contexts.

## Introduction

Adolescent substance use has emerged as a critical global public health challenge, with a notable surge observed in developing countries (1). The consumption of alcohol, tobacco products, and illicit substances is a widespread phenomenon across all demographics, with adolescents exhibiting particularly high prevalence rates (2). Defined as any chemical compound that, upon administration or ingestion, alters psychological processes including perception, consciousness, cognition, mood, and emotions (3), these substances pose significant risks to adolescent health and well-being.

Substance use, defined as the non-medical and non-scientific use of controlled psychoactive substances (4), was historically associated with urban environments. However, recent research indicates a shift in this pattern. Substance use, including illicit drugs and alcohol, is now observed across diverse socioeconomic and educational backgrounds, including school-aged children (5,6), contradicting previous assumptions.

Evolving societal and economic factors have contributed to a rise in substance use, with notable trends including polydrug use and increased tobacco consumption among adolescent girls (7). These findings highlight the need for updated public health strategies. Substance abuse, encompassing the excessive consumption of illicit drugs, prescription and over-the-counter medications, and alcohol, poses a significant global health challenge. Recent data reveals a concerning rise in drug use worldwide, with 275 million users reported in the past year, a 22% increase from 2010. Projections indicate an 11% global increase by 2030, with Africa expected to experience a disproportionate 40% surge. In 2019 alone, drug use resulted in approximately half a million deaths and the loss of 18 million years of healthy life (8).

Adolescents are particularly vulnerable to substance abuse, with studies in Africa highlighting a high prevalence linked to various physical and psychosocial issues. This vulnerability is exacerbated by the fact that adolescence is a critical period of brain development, making young people more susceptible to the detrimental effects of these substances (9–11).

Alcohol consumption is another major concern, contributing to 3 million deaths annually and a substantial global disease burden (12). In Sub-Saharan Africa, tobacco and alcohol are the most prevalent substances used by adolescents, with an overall substance use prevalence of 41.6% (Jumbe et al., 2021). Tobacco use alone claims over 8 million lives each year, making it a leading cause of preventable death worldwide (12). The COVID-19 pandemic has further amplified the issue of substance abuse, particularly among young people (13). Increased stress, isolation, and disruptions to daily life have contributed to a rise in substance use as a coping mechanism (7).

Substance abuse has far-reaching negative consequences, including impaired mental function, increased risk of mental health disorders and dependence, unemployment, cancers, and other health problems (14). These effects extend to families and communities, causing psychological distress, economic strain, and increased healthcare costs. Given the existing burden of disease and socio-political challenges in Sub-Saharan Africa, the escalating substance abuse crisis demands urgent attention. It is imperative to address this issue before it reaches a critical point, as the consequences for individuals, families, and society are devastating.

The global prevalence of substance use and abuse is on the rise, with the World Drug Report projecting an 11% increase in drug users by 2030 due to demographic shifts, and it is expected to be more pronounced in Africa, where the younger population, more prone to drug use, is proportionally larger (United Nations Office on Drugs and Crime, 2021). Substance abuse significantly contributes to the loss of disability-adjusted life years (DALYs) in young people worldwide, with African rates 2.5 times higher than in high-income countries (Castelpietra et al., 2022). While research on substance use is established, evolving trends such as experimentation with mixing substances, volatile inhalant use, and the discovery of novel intoxicants necessitate ongoing investigation into this issue (Ogundipe et al., 2018).

Ghana has not been immune to the escalating problem of substance use. There is a concerning increase in drug trafficking and consumption in the region over the past decade, impacting security, governance, and development (15). In recent years, Ghana, like other African nations, has witnessed a rise in substance use among youth, including tobacco, alcohol, and marijuana (11,14,16). The 2012 Global School-Based Student Health Survey (GSHS) found that 8.3% of students smoked cigarettes, 15.4% consumed alcohol, and 9.4% reported past-month intoxication, with 94.3% of smokers initiating before age 14 (17).

Substance use is particularly prevalent in Senior High Schools (18,19). While cigarette smoking remains the most common form of tobacco use, waterpipe tobacco smoking (WTS), or shisha, is increasingly popular among adolescents, particularly females, due to misconceptions about its harmfulness compared to cigarettes (20,21). The consequences of substance abuse are severe, ranging from immune system suppression and infectious diseases to cardiovascular complications and mortality.

Research studies have found a correlation between energy drink consumption and substance use among adolescents, including alcohol, tobacco, cannabis, and non-medical prescription drugs (22). In response, the Food and Drugs Authority (FDA) has launched the “DAABI CAMPAIGN” aimed at raising awareness about substance abuse in SHS (21).

Intrapersonal, interpersonal, and societal factors can all contribute to adolescent substance abuse. Intrapersonal factors, such as knowledge and perception of substance use risks and self-esteem, significantly influence adolescent engagement with substances (23). Interpersonal relationships, especially with peers and family members who exhibit substance use patterns, are a critical risk factor for adolescents (24,25). Specifically, strained parent-child relationships, especially for girls, are associated with increased alcohol and marijuana use (26,27).

Social factors, such as peer influence, family history of substance abuse, and community norms, play a substantial role in adolescent substance use initiation and escalation (25). Research indicates a strong correlation between adolescent alcohol use and the alcohol use patterns of their peers (17). Furthermore, parental substance abuse is a significant risk factor for adolescent substance use, contributing to exposure to adverse childhood experiences and increasing the likelihood of developing substance use issues (28).

The accessibility and availability of substances within the community, influenced by local and national regulations and policies, can either facilitate or inhibit adolescent substance use (29). Furthermore, media representations of substance use also significantly impact adolescents, serving as a primary source of information and potentially normalizing substance use behaviours (30,31).

Several additional factors can increase adolescent vulnerability to substance use, including experimentation, lack of knowledge, inadequate parental supervision, peer and family pressure, gang involvement, mental health issues, truancy, and academic difficulties (25). Adolescents may initiate substance use due to a variety of reasons, such as stress relief, curiosity, or lack of awareness regarding the consequences for themselves and others. Strained family relationships and poor communication can also contribute to adolescent dissatisfaction and substance use as a coping mechanism (28).

Previous research in Ghana has documented the use of alcohol, tobacco, and other substances among adolescents, but there remains a need for updated, context-specific data to inform effective prevention and intervention strategies. Accra’s status as the national educational and economic centre means that findings from this study have the potential to inform policy and practice not only within the city but also across the country.

By focusing on in-school adolescents in Accra, this study aims to shed light on the current patterns of substance use, identify the key correlates and risk factors, and contribute to a deeper understanding of the complex interplay of individual, social, and environmental factors influencing substance use among this vulnerable population.

This study seeks to examine the correlates of substance use among SHS students and examine current trends and patterns in relation to socio-demographic and psycho-social circumstances. This knowledge is crucial for informing comprehensive interventions and treatment programmes for adolescents.

## Methods

### Study Design

A quantitative cross-sectional survey was conducted among Senior High School (SHS) students within the Accra Metropolitan Area (AMA), Greater Accra Region, Ghana. The AMA is one of 26 Metropolitan, Municipal, and District Assemblies (MMDAs) within the Greater Accra Region (GAR). Educational oversight within the AMA falls under the jurisdiction of the Accra Metropolitan Education Office (AMEO) of the Ghana Education Service (GES).

Accra, the capital of Ghana and the site of this study, presents a dynamic and complex landscape for examining substance use among in-school adolescents. As a rapidly urbanizing city, Accra grapples with social, economic, and educational disparities that may influence adolescent behaviour. The city’s diverse population includes various ethnic groups (Ga, Akan, Ewe), religious affiliations (Christian, Muslim, Traditional), and socioeconomic levels, creating a multifaceted context for substance use patterns.

The Ghanaian education system in Accra consists of public and private schools with varying levels of resources and quality. While education is highly valued in Ghanaian culture, challenges such as overcrowding, limited infrastructure, and varying teaching standards persist. These factors, coupled with the pressures of academic expectations and social influences, may contribute to the vulnerability of some adolescents to substance use.

### Population and sample

The study population consisted of students attending public Senior High Schools (SHS) within the Accra Metropolitan Area (AMA). Inclusion criteria encompassed all SHS Form 1 and 2 students present on the day of data collection who expressed willingness to participate. Exclusion criteria included students who were unwell or absent from school on the data collection day. Two public schools were randomly selected from a total of five eligible institutions to participate in the study. Sample size determination was calculated utilizing Cochran’s formula for categorical data:

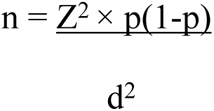

Where p = proportion of substance use among adolescents 45.6% (32)

z^2^= 95% confidence interval corresponding to the value of 1.96

d^2^ = proportion of sampling error tolerated at 0.05% (to increase the accuracy).

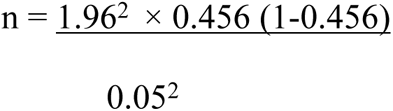

n = 381.19

Assuming a non-response rate of about 10%, 382 × 0.1 = 38.2, the actual sample size is 382 + 39 = 421. Therefore, the minimum sample size was 421. This methodology ensured a representative sample of the target population, allowing for generalizable conclusions regarding the research objectives.

Participant selection was achieved through a multi-stage sampling approach.

Initially, two schools were randomly chosen from a pool of five potential institutions. Subsequently, within each selected school, classes were randomly selected from the comprehensive list of available classes. Finally, a systematic sampling method was employed within each chosen class to identify the requisite number of student participants. Data collection was limited to students in Senior High School (SHS) year 1 and year 2, as SHS 3 students had already completed their academic year at the time of data collection. Data collection took place from 12^th^ January 2023 to 23^rd^ February 2023.

### Measures

A structured questionnaire consisting of several sections was designed to address the research objectives. The first section gathered demographic information, including age, gender, socioeconomic status, and educational level, to characterize the sample population.

The second section explored the availability and accessibility of five substances prevalent in the region: alcohol, cigarettes, shisha, and marijuana (wee)f. Participants indicated whether they had encountered each substance in their environment and assessed the ease of obtaining each on a five-point Likert scale ranging from “very easy” to “very difficult.” This section also investigated the primary sources from which adolescents obtained these substances, such as friends, family members, or illicit vendors. Additionally, participants reported the contexts in which they were most likely to use these substances, including social gatherings, after-school hours, or during periods of stress.

The third section delved into the assessment of substance use patterns and potential dependence among the adolescent participants. The CAGE-AID Substance Abuse Screening Tool, a validated instrument comprising questions related to alcohol and drug use, was employed for this purpose. The tool serves as an initial screening mechanism, identifying individuals who may require further evaluation for substance use disorders. Item responses were scored dichotomously, with a cumulative score of two or greater suggesting a potential need for intervention.

In addition to substance use, the study also examined self-esteem as a potential contributing factor to substance use behaviours. The Rosenborg Self-esteem Scale, a widely used 10-item uni-dimensional scale, was administered to assess participants’ global self-worth. The scale captures both positive and negative feelings about oneself, providing a comprehensive evaluation of self-esteem levels. Responses were recorded on a four-point Likert scale and subsequently scored using a combined rating method, where higher scores signified greater self-esteem.

### Statistical analysis

Data were exported to Stata version 16.1 for cleaning, processing, and analysis. A two-pronged analytical approach was employed, incorporating both difference-in-proportions and inferential analyses. To assess differences in proportions, Pearson’s chi-square (χ2) test was utilized, with Fisher’s exact test substituted in cases where any 2 × 2 table cell frequency was less than 5%. Inferential analysis was conducted via logistic regression, employing both univariate and multivariate approaches. Regression models were estimated using a 95% confidence interval, with a p-value < 0.05 denoting statistical significance. This sequential approach allowed for initial identification of significant proportional differences, followed by quantification of the association strength through logistic regression. Substance use dependence was assessed descriptively using the CAGE–AID Substance Abuse Screening Tool. A “yes” response to two or more of the four CAGE-AID questions was indicative of potential substance dependence.

### Ethical considerations

Ethical clearance for this study was obtained from the Ghana Health Service Ethics Review Committee (GHS-ERC: 025/09/22). Furthermore, permission was granted by the Accra Metro Education Office of the Ghana Education Service and the respective heads of the Senior High Schools involved in participant recruitment. For participants under the age of 18, both written parental consent and participant assent were prerequisites for study enrolment. All participants received thorough information regarding the study’s objectives, procedures, and potential risks and benefits, enabling informed decision-making about their participation. The study adhered to strict anonymity, confidentiality, and privacy protocols. All adolescent participants provided voluntary informed consent.

## Results

### Socio-demographic characteristics of participants

A total of 425 adolescents and young adults participated in the study. Participants were primarily female, with a slight majority in SHS 1. Most participants identified as Christian, while a minority identified as Muslim. Living arrangements were diverse, with participants living with their mothers, both parents, both parents and siblings, relatives, or alone. A notable proportion of participants reported working while still attending school. Demographic details of the sample are presented in Table 1.

**Table 1.**
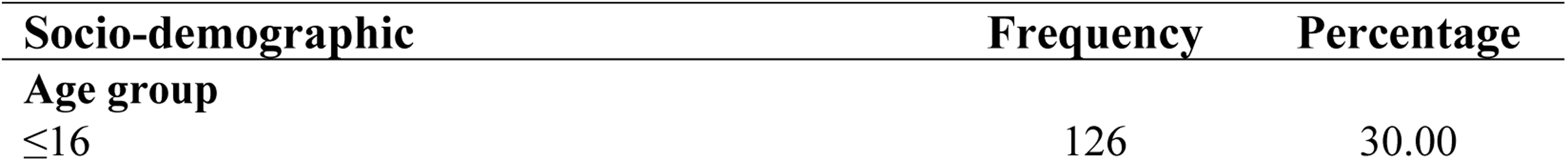
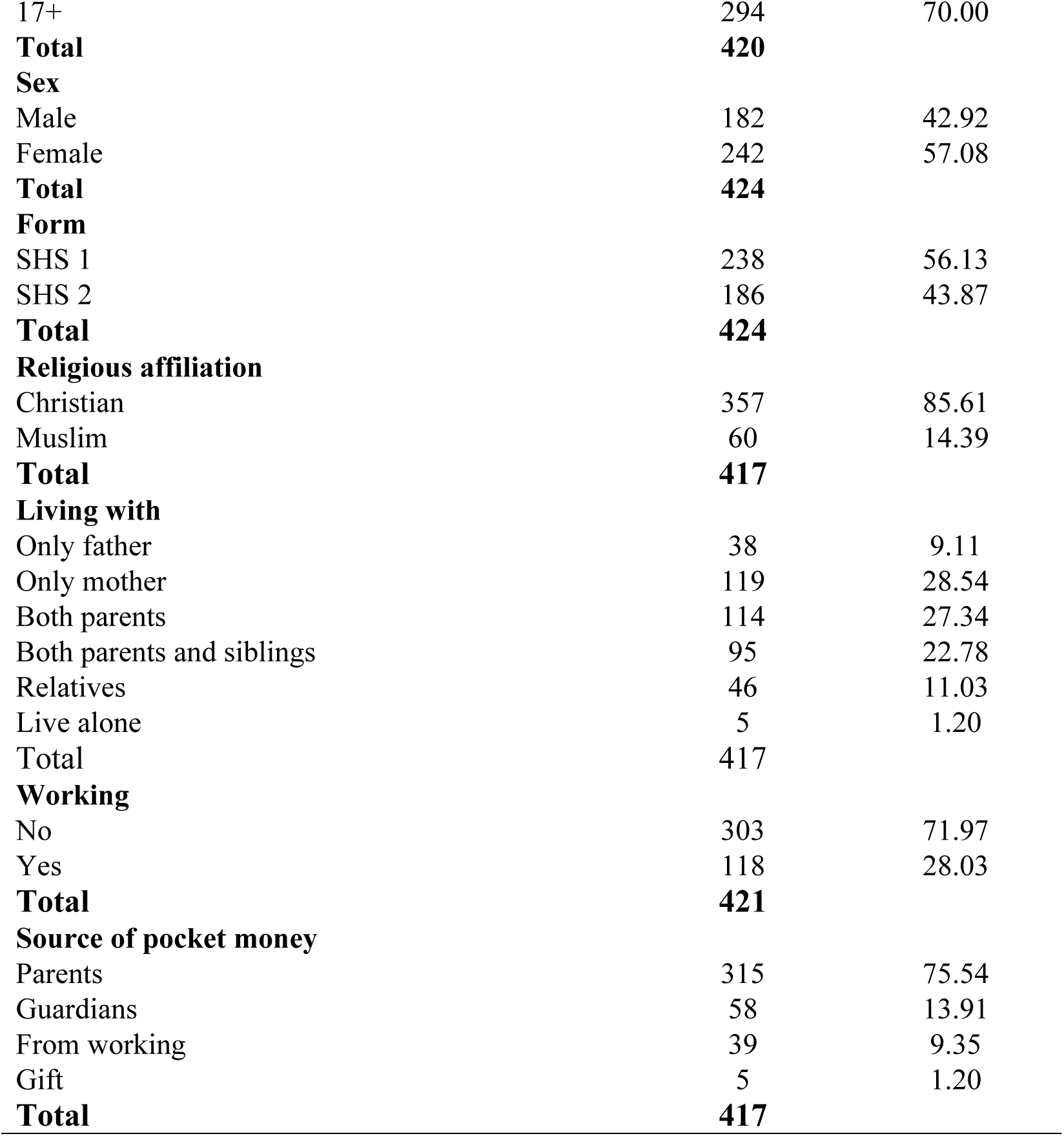
Socio-demographic characteristics of participants.

### Types and sources of substances used

The primary source of all substances for participants was friends. This was followed by drug dealers, then online retailers and markets, with schools being the least common source. For alcohol specifically, friends were the most frequent source, followed by drug peddlers, online retailers, and markets, with schools being the least common. Shisha was primarily obtained from friends, with drug peddlers as a secondary source. Marijuana users also mainly sourced their supply from friends, followed by drug peddlers and markets. Cigarettes were primarily obtained from friends as well. Table 2 contains detailed statistics on the frequency of each source.

**Table 2.** Types and sources of substances used.

| Type of Substance | Source of substance use |  |  |  |  | Total |
| --- | --- | --- | --- | --- | --- | --- |
|  | School | Friends | Online/social media | Market | Drug peddlers |  |
|  | n(%) | n(%) | n(%) | n(%) | n(%) | n |
| Alcohol (n=79) | 17(21.52) | 66(83.54) | 32(40.51) | 30(37.97) | 57(72.15) | **202** |
| Cigarette (n=16) | 3(20.00) | 12(80.00) | 5(33.33) | 4(26.67) | 8(53.33) | **32** |
| Shisha (n=51) | 9(17.65) | 45(88.24) | 17(33.33) | 13(25.49) | 30(58.82) | **114** |
| Marijuana (n=30) | 3(10.00) | 24(80.00) | 5(16.67) | 9(30.00) | 20(66.67) | **61** |
| **Total** | **32(18.18)** | **147(83.52)** | **59(33.52)** | **56(31.82)** | **115(65.34)** | **409** |

### Personal characteristics, social and intrapersonal factors, and alcohol use

The prevalence of alcohol use among participants was 19.4% (n=79). Univariate analysis identified several factors potentially associated with alcohol use. However, multivariate logistic regression (Table 3) revealed that only sex, living situation, and peer influence remained significantly associated with alcohol use.

**Table 3.** Personal characteristics, social and intrapersonal factors, and alcohol use.

| Socio-demographic | Alcohol use | | $\chi^2$(p-value) | Logistic Regression | |
| --- | --- | --- | --- | --- | --- |
|  | No=336(80.96) | Yes=79(19.04) |  | cOR[95%CI]p-value | aOR[95%CI]p-value |
|  | n(%) | n(%) |  |  |  |
| **Age group (n=411)** |  |  |  |  |  |
| ≤16 | 102(82.26) | 22(17.74) | 0.25(0.617) | **ref** | **ref** |
| 17+ | 230(80.14) | 57(19.86) |  | 1.15[0.67-1.98]0.617 | 1.10[0.60-2.01]0.769 |
| **Sex (n=414)** |  |  |  |  |  |
| Female | 203(85.65) | 34(14.35) | 8.05(0.005)** | **ref** | **ref** |
| Male | 132(74.58) | 45(25.42) |  | 2.04[1.24-3.35]0.005** | 1.88[1.04-3.40]0.036* |
| **Form (n=414)** |  |  |  |  |  |
| SHS 2 | 156(86.67) | 24(13.33) | 6.32(0.012)* | **ref** | **ref** |
| SHS 1 | 180(76.92) | 54(23.08) |  | 1.95[1.15-3.30]0.013* | 1.75[0.97-3.14]0.063 |
| **Religious affiliation (n=408)** |  |  |  |  |  |
| Muslim | 53(88.33) | 7(11.67) | 2.53(0.112) | **ref** | **ref** |
| Christian | 277(79.6) | 71(20.4) |  | 1.94[0.85-4.46]0.118 | 1.34[0.55-3.23]0.517 |
| **Whom do you live with (n=408)** |  |  |  |  |  |
| Only father | 29(76.32) | 9(23.68) | 11.61(0.040)* | **ref** | **ref** |
| Only mother | 85(74.56) | 29(25.44) |  | 1.10[0.47-2.60]0.829 | 0.92[0.38-2.25]0.862 |
| Both parents | 98(85.96) | 16(14.04) |  | 0.53[0.21-1.32]0.170 | 0.53[0.20-1.37]0.188 |
| Both parents and siblings | 72(79.12) | 19(20.88) |  | 0.85[0.34-2.10]0.725 | 0.82[0.31-2.14]0.680 |
| Relatives | 43(93.48) | 3(6.52) |  | 0.22[0.06-0.90]0.035* | 0.22[0.05-0.95]0.043* |
| Live alone | 5(100) | 0(0.0) |  | - | - |
| **Working apart from schooling (n=412)** |  |  |  |  |  |
| No | 246(82.83) | 51(17.17) | 2.75(0.097) | **ref** | **ref** |
| Yes | 87(75.65) | 28(24.35) |  | 1.55[0.92-2.62]0.099 | 1.00[0.49-2.06]0.999 |
| **Where participant get pocket money from? (n=408)** |  |  |  |  |  |
| Parents | 253(82.14) | 55(17.86) | 8.28(0.041)* | **ref** | **ref** |
| Guardians | 49(85.96) | 8(14.04) |  | 0.75[0.34-1.68]0.485 | 1.00[0.41-2.47]0.993 |
| From working | 25(65.79) | 13(34.21) |  | 2.39[1.15-4.97]0.019* | 2.06[0.78-5.41]0.143 |
| Gift | 3(60) | 2(40) |  | 3.07[0.50-18.83]0.226 | 4.57[0.37-56.13]0.235 |
| **Exposure to substance by friends (n=415)** |  |  |  |  |  |
| No | 101(90.99) | 10(9.01) | 9.88(0.002)** | **ref** | **ref** |
| Yes | 235(77.3) | 69(22.7) |  | 2.97[1.47-5.99]0.002** | 2.34[1.03-5.32]0.043* |
| **Exposure to substance by family (n=415)** |  |  |  |  |  |
| No | 147(88.02) | 20(11.98) |  | **ref** | **ref** |
| Yes | 189(76.21) | 59(23.79) | 9.04(0.003)** | 2.29[1.32-3.98]0.003** | 1.78[0.95-3.35]0.073 |
| **Social media exposure to substance (n=415)** |  |  |  |  |  |
| Yes | 139(81.29) | 32(18.71) |  | **ref** | **ref** |
| No | 197(80.74) | 47(19.26) | 0.02(0.889) | 1.04[0.63-1.71]0.889 | 1.13[0.64-2.01]0.671 |
| **level of access to substance use (n=415)** |  |  |  |  |  |
| High access | 189(81.12) | 44(18.88) |  | **ref** | **ref** |
| Low access | 147(80.77) | 35(19.23) | 0.008(0.929) | 1.02[0.62-1.68]0.929 | 1.07[0.60-1.91]0.813 |
| **Self-esteem (n=415)** |  |  |  |  |  |
| High esteem | 149(81.87) | 33(18.13) |  | **ref** | **ref** |
| Low esteem | 183(79.91) | 46(20.09) | 1.20(0.548) | 1.17[0.71-1.91]0.546 | 0.90[0.51-1.60]0.724 |
| **Perception on substance use (n=415)** |  |  |  |  |  |
| High | 89(82.41) | 19(17.59) |  | **ref** | **ref** |
| Low | 152(79.58) | 39(20.42) | 0.45(0.799) | 1.20[0.65-2.21]0.554 | 1.25[0.62-2.51]0.538 |
| Average | 95(81.9) | 21(18.1) |  | 1.04[0.52-2.06]0.921 | 1.04[0.47-2.28]0.923 |

Note: Abbreviation: n=total number of samples involved as connected to a particular variable; cOR= crude odds ratio; aOR= adjusted odds ratio; CI=confidence interval; \*p<0.05; \*\*p<0.01

Specifically, males were 1.88 times more likely to use alcohol than females (AOR=1.88, 95% CI: 1.04-3.40, p=0.036). Living with relatives was associated with a decreased likelihood of alcohol use (AOR=0.22, 95% CI: 0.05-0.95, p=0.043), while having friends who use alcohol increased the likelihood of use by 2.34 times (AOR=2.34, 95% CI: 1.03-5.32, p=0.043).

### Personal characteristics, social and intrapersonal factors, and cigarette use

The prevalence of cigarette smoking among the study population was 3.77% (n=15). Neither age group (AOR=0.70, 95% CI: 0.19-2.61, p=0.593) nor sex (AOR=1.14, 95% CI: 0.30-4.41, p=0.847) were significantly associated with smoking status (Table 4). Univariate analysis indicated that participants who worked while attending school were more likely to smoke (COR=3.11; 95% CI: 1.10-8.82, p=0.032). However, this association was not statistically significant in the adjusted model. Exposure to cigarette use by a family member was significantly associated with smoking (AOR=13.23, 95% CI: 1.63-107.35, p=0.016).

**Table 4.** Personal characteristics, social and intrapersonal factors, and cigarette use.

| Socio-demographic | Cigarette use | | $\chi^2$(p-value) | Logistic regression | |
| --- | --- | --- | --- | --- | --- |
|  | No=383(96.23) | Yes=15(3.77) |  | cOR[95%CI] p-value | aOR[95%CI] p-value |
|  | n(%) | n(%) |  |  |  |
| **Age group (n=394)** |  |  |  |  |  |
| ≤16 | 116(96.67) | 4(3.33) | 0.11(0.745) | **ref** | **ref** |
| 17+ | 263(95.99) | 11(4.01) |  | 1.21[0.38-3.89]0.746 | 0.70[0.19-2.61]0.593 |
| **Sex (n=397)** |  |  |  |  |  |
| Female | 224(96.97) | 7(3.03) | 0.85(0.356) | **ref** | **ref** |
| Male | 158(95.18) | 8(4.82) |  | 1.62[0.58-4.57]0.361 | 1.14[0.30-4.41]0.847 |
| **Form (n=397)** |  |  |  |  |  |
| SHS 2 | 170(96.59) | 6(3.41) | 0.12(0.731) | **ref** | **ref** |
| SHS 1 | 212(95.93) | 9(4.07) |  | 1.20[0.42-3.45]0.731 | 1.19[0.33-4.25]0.789 |
| **Religious affiliation (n=391)** |  |  |  |  |  |
| Muslim | 57(96.61) | 2(3.39) | 0.007(0.932) | **ref** | **ref** |
| Christian | 320(96.39) | 12(3.61) |  | 1.07[0.23-4.91]0.932 | 0.82[0.16-4.33]0.818 |
| **Whom do you live with (n=392)** |  |  |  |  |  |
| Only father | 35(94.59) | 2(5.41) | 6.22(0.285) | **ref** | **ref** |
| Only mother | 103(94.5) | 6(5.5) |  | 1.02[0.20-5.30]0.982 | 0.62[0.12-3.19]0.563 |
| Both parents | 99(94.29) | 6(5.71) |  | 1.06[0.20-5.52]0.944 | 0.81[0.15-4.32]0.809 |
| Both parents and siblings | 90(100) | 0(0.0) |  | - | - |
| Relatives | 45(97.83) | 1(2.17) |  | 0.39[0.03-4.48]0.449 | 0.37[0.01-15.60]0.603 |
| Live alone | 5(100) | 0(0.0) |  | - | - |
| **Working apart from schooling (n=395)** |  |  |  |  |  |
| No | 278(97.54) | 7(2.46) | 5.04(0.025)* | **ref** | **ref** |
| Yes | 102(92.73) | 8(7.27) |  | 3.11[1.10-8.82]0.032* | 3.36[0.72-15.68]0.123 |
| **Where participant get pocket money from? (n=391)** |  |  |  |  |  |
| Parents | 282(96.25) | 11(3.75) | 2.85(0.416) | **ref** | **ref** |
| Guardians | 56(98.25) | 1(1.75) |  | 0.46[0.06-3.63]0.459 | 0.29[0.04-2.06]0.214 |
| From working | 33(91.67) | 3(8.33) |  | 2.33[0.62-8.80]0.212 | 0.93[0.21-4.03]0.920 |
| Gift | 5(100) | 0(0.0) |  | - | - |
| **Exposure to substance by friends (n=398)** |  |  |  |  |  |
| No | 105(97.22) | 3(2.78) | 0.40(0.526) | **ref** | **ref** |
| Yes | 278(95.86) | 12(4.14) |  | 1.51[0.42-5.47]0.53 | 1.33[0.23-7.62]0.749 |
| **Exposure to substance by family (n=398)** |  |  |  |  |  |
| No | 158(98.75) | 2(1.25) |  | **ref** | **ref** |
| Yes | 225(94.54) | 13(5.46) | 4.68(0.031)* | 4.56[1.01-20.55]0.048* | 13.23[1.63-107.35]0.016* |
| **Social media exposure to substance (n=398)** |  |  |  |  |  |
| Yes | 160(96.97) | 5(3.03) | 0.42(0.515) | **ref** | **ref** |
| No | 223(95.71) | 10(4.29) |  | 1.43[0.48-4.28]0.518 | 1.49[0.40-5.50]0.554 |
| **Level of access to substance use (n=398)** |  |  |  |  |  |
| High access | 219(97.77) | 5(2.23) | 3.34(0.068) | **ref** | **ref** |
| Low access | 164(94.25) | 10(5.75) |  | 2.67[0.89-7.97]0.078 | 2.23[0.67-7.47]0.192 |
| **Self-esteem (n=398)** |  |  |  |  |  |
| High esteem | 171(97.16) | 5(2.84) | 0.98(0.614) | **ref** | **ref** |
| Low esteem | 208(95.41) | 10(4.59) |  | 1.68[0.56-5.02]0.351 | 0.83[0.21-3.32]0.794 |
| **Perception on substance use (n=398)** |  |  |  |  |  |
| High | 100(99.01) | 1(0.99) |  | **ref** | **ref** |
| Low | 175(95.11) | 9(4.89) | 2.92(0.232) | 5.14[0.64-41.30]0.123 | 4.96[0.42-58.05]0.202 |
| Average | 108(95.58) | 5(4.42) |  | 4.63[0.53-40.42]0.166 | 3.95[0.30-51.73]0.295 |

Note: Abbreviation: n=total number of samples involved as connected to a particular variable; cOR= crude odds ratio; aOR= adjusted odds ratio; CI=confidence interval; \*p<0.05

### Personal characteristics, social and intrapersonal factors, and shisha use

The prevalence of shisha use among participants was 12.59% (n=51). Several factors were found to be significantly associated with shisha use, including age, employment status while attending school, source of pocket money, and shisha use among family and friends (p<0.01).

Univariate analyses revealed significant associations between several factors and shisha use (Table 5). Participants who earned their pocket money through work were three times more likely to use shisha than those who received pocket money from parents (COR=3.32, 95% CI: 1.46-7.53, p=0.004). Additionally, respondents with a family member who used shisha were more than twice as likely to use shisha themselves (COR=2.39, 95% CI: 1.21-4.72, p=0.012).

**Table 5.**
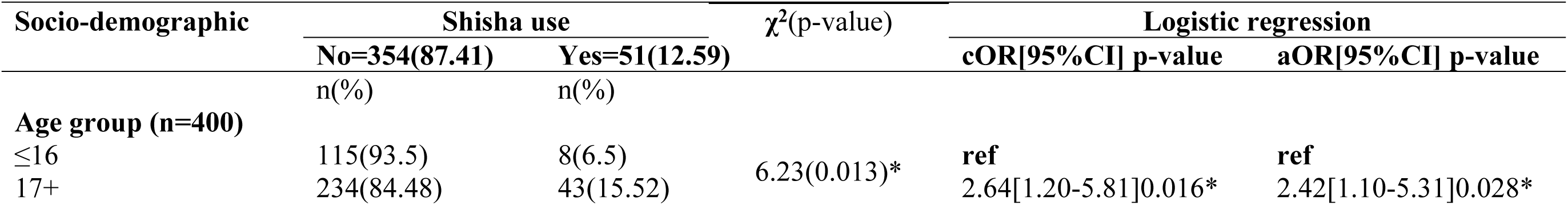
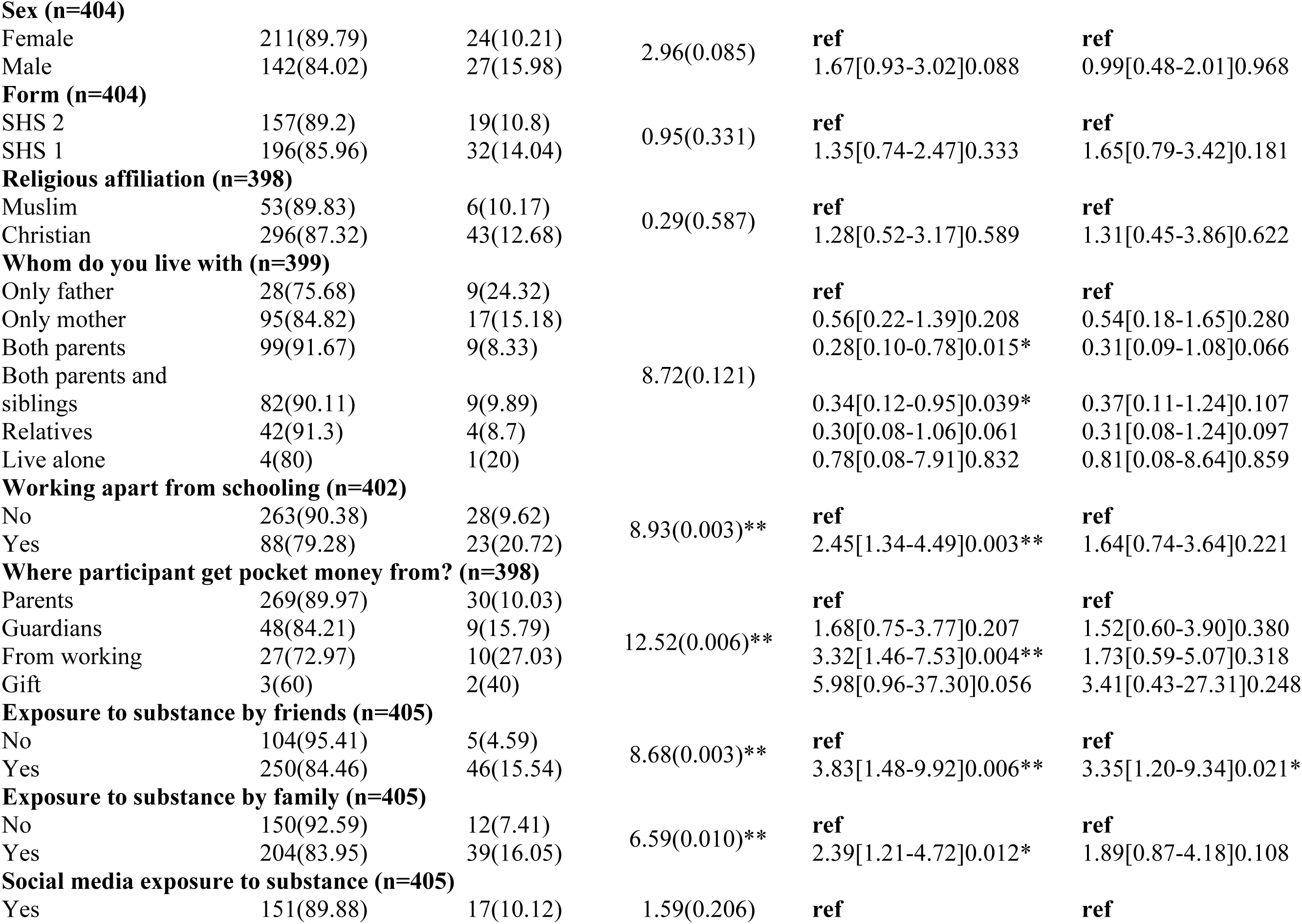
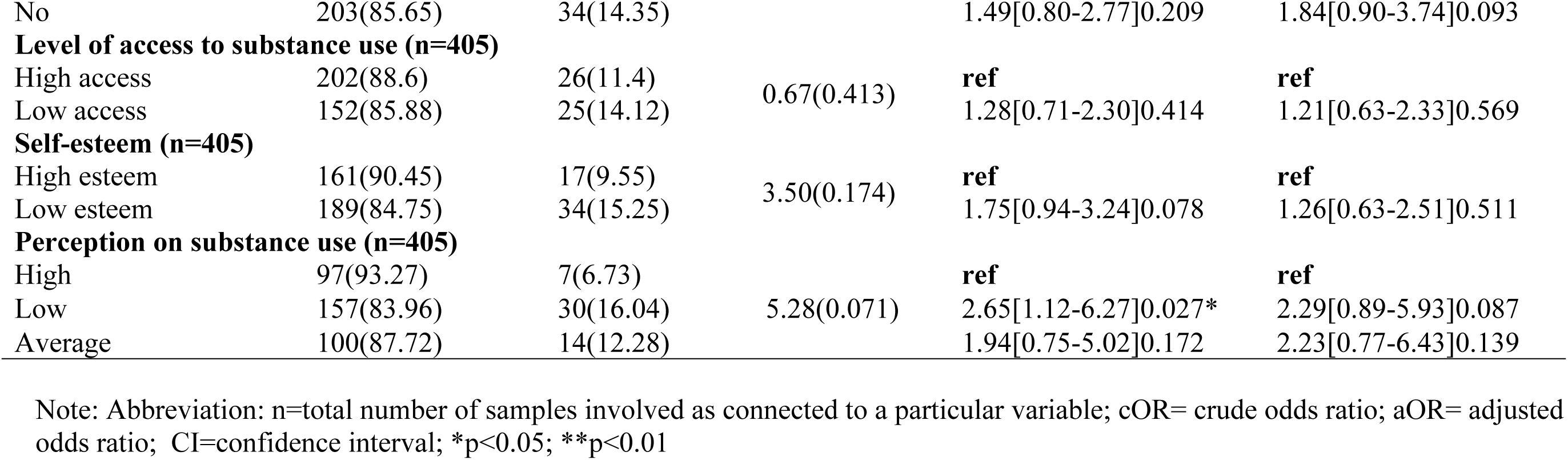
Personal characteristics, social and intrapersonal factors, and shisha use.

| Socio-demographic | Shisha use | | $\chi^2$(p-value) | Logistic regression | |
| --- | --- | --- | --- | --- | --- |
|  | No=354(87.41) | Yes=51(12.59) |  | cOR[95%CI] p-value | aOR[95%CI] p-value |
|  | n(%) | n(%) |  |  |  |
| **Age group (n=400)** |  |  |  |  |  |
| ≤16 | 115(93.5) | 8(6.5) | 6.23(0.013)* | **ref** | **ref** |
| 17+ | 234(84.48) | 43(15.52) |  | 2.64[1.20-5.81]0.016* | 2.42[1.10-5.31]0.028* |
| **Sex (n=404)** |  |  |  |  |  |
| Female | 211(89.79) | 24(10.21) | 2.96(0.085) | **ref** | **ref** |
| Male | 142(84.02) | 27(15.98) |  | 1.67[0.93-3.02]0.088 | 0.99[0.48-2.01]0.968 |
| **Form (n=404)** |  |  |  |  |  |
| SHS 2 | 157(89.2) | 19(10.8) | 0.95(0.331) | **ref** | **ref** |
| SHS 1 | 196(85.96) | 32(14.04) |  | 1.35[0.74-2.47]0.333 | 1.65[0.79-3.42]0.181 |
| **Religious affiliation (n=398)** |  |  |  |  |  |
| Muslim | 53(89.83) | 6(10.17) | 0.29(0.587) | **ref** | **ref** |
| Christian | 296(87.32) | 43(12.68) |  | 1.28[0.52-3.17]0.589 | 1.31[0.45-3.86]0.622 |
| **Whom do you live with (n=399)** |  |  |  |  |  |
| Only father | 28(75.68) | 9(24.32) | 8.72(0.121) | **ref** | **ref** |
| Only mother | 95(84.82) | 17(15.18) |  | 0.56[0.22-1.39]0.208 | 0.54[0.18-1.65]0.280 |
| Both parents | 99(91.67) | 9(8.33) |  | 0.28[0.10-0.78]0.015* | 0.31[0.09-1.08]0.066 |
| Both parents and siblings | 82(90.11) | 9(9.89) |  | 0.34[0.12-0.95]0.039* | 0.37[0.11-1.24]0.107 |
| Relatives | 42(91.3) | 4(8.7) |  | 0.30[0.08-1.06]0.061 | 0.31[0.08-1.24]0.097 |
| Live alone | 4(80) | 1(20) |  | 0.78[0.08-7.91]0.832 | 0.81[0.08-8.64]0.859 |
| **Working apart from schooling (n=402)** |  |  |  |  |  |
| No | 263(90.38) | 28(9.62) | 8.93(0.003)** | **ref** | **ref** |
| Yes | 88(79.28) | 23(20.72) |  | 2.45[1.34-4.49]0.003** | 1.64[0.74-3.64]0.221 |
| **Where participant get pocket money from? (n=398)** |  |  |  |  |  |
| Parents | 269(89.97) | 30(10.03) | 12.52(0.006)** | **ref** | **ref** |
| Guardians | 48(84.21) | 9(15.79) |  | 1.68[0.75-3.77]0.207 | 1.52[0.60-3.90]0.380 |
| From working | 27(72.97) | 10(27.03) |  | 3.32[1.46-7.53]0.004** | 1.73[0.59-5.07]0.318 |
| Gift | 3(60) | 2(40) |  | 5.98[0.96-37.30]0.056 | 3.41[0.43-27.31]0.248 |
| **Exposure to substance by friends (n=405)** |  |  |  |  |  |
| No | 104(95.41) | 5(4.59) | 8.68(0.003)** | **ref** | **ref** |
| Yes | 250(84.46) | 46(15.54) |  | 3.83[1.48-9.92]0.006** | 3.35[1.20-9.34]0.021* |
| **Exposure to substance by family (n=405)** |  |  |  |  |  |
| No | 150(92.59) | 12(7.41) | 6.59(0.010)** | **ref** | **ref** |
| Yes | 204(83.95) | 39(16.05) |  | 2.39[1.21-4.72]0.012* | 1.89[0.87-4.18]0.108 |
| **Social media exposure to substance (n=405)** |  |  |  |  |  |
| Yes | 151(89.88) | 17(10.12) | 1.59(0.206) | **ref** | **ref** |
| No | 203(85.65) | 34(14.35) |  | 1.49[0.80-2.77]0.209 | 1.84[0.90-3.74]0.093 |
| **Level of access to substance use (n=405)** |  |  |  |  |  |
| High access | 202(88.6) | 26(11.4) | 0.67(0.413) | **ref** | **ref** |
| Low access | 152(85.88) | 25(14.12) |  | 1.28[0.71-2.30]0.414 | 1.21[0.63-2.33]0.569 |
| **Self-esteem (n=405)** |  |  |  |  |  |
| High esteem | 161(90.45) | 17(9.55) | 3.50(0.174) | **ref** | **ref** |
| Low esteem | 189(84.75) | 34(15.25) |  | 1.75[0.94-3.24]0.078 | 1.26[0.63-2.51]0.511 |
| **Perception on substance use (n=405)** |  |  |  |  |  |
| High | 97(93.27) | 7(6.73) | 5.28(0.071) | **ref** | **ref** |
| Low | 157(83.96) | 30(16.04) |  | 2.65[1.12-6.27]0.027* | 2.29[0.89-5.93]0.087 |
| Average | 100(87.72) | 14(12.28) |  | 1.94[0.75-5.02]0.172 | 2.23[0.77-6.43]0.139 |

Note: Abbreviation: n=total number of samples involved as connected to a particular variable; cOR= crude odds ratio; aOR= adjusted odds ratio; CI=confidence interval; \*p<0.05; \*\*p<0.01

Participants with a low perception of the consequences of shisha use were also more likely to use shisha compared to those with a higher perception (COR=2.65, 95% CI: 1.12-6.27, p=0.027). Age was another significant factor, with participants aged 17 years and above being more likely to use shisha than those aged 16 and below (AOR=2.42, 95% CI: 1.10-5.31, p=0.028). Finally, respondents with friends who smoked shisha were three times more likely to use it themselves (AOR=3.35, 95% CI: 1.20-9.34, p=0.021).

### Personal characteristics, social and intrapersonal factors, and marijuana use

The prevalence of the use of marijuana among participants was 30 (7.5%). From the multivariate analysis, factors associated with marijuana use include family exposure (AOR=3.66, 95% CI: 1.21-11.03, p=0.021) and non-exposure to social media (AOR=5.80, 95% CI:1.84-18.28, p=0.003). Respondents who live with parents and siblings (AOR=0.14, 95% CI: 0.03-0.60, P=0.008) were less likely to use marijuana as opposed to those who live with only their father (Table 6).

**Table 6.**
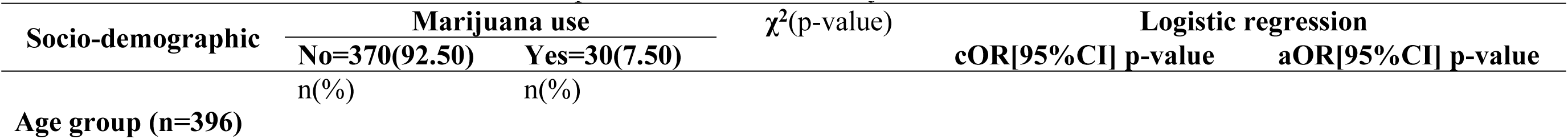
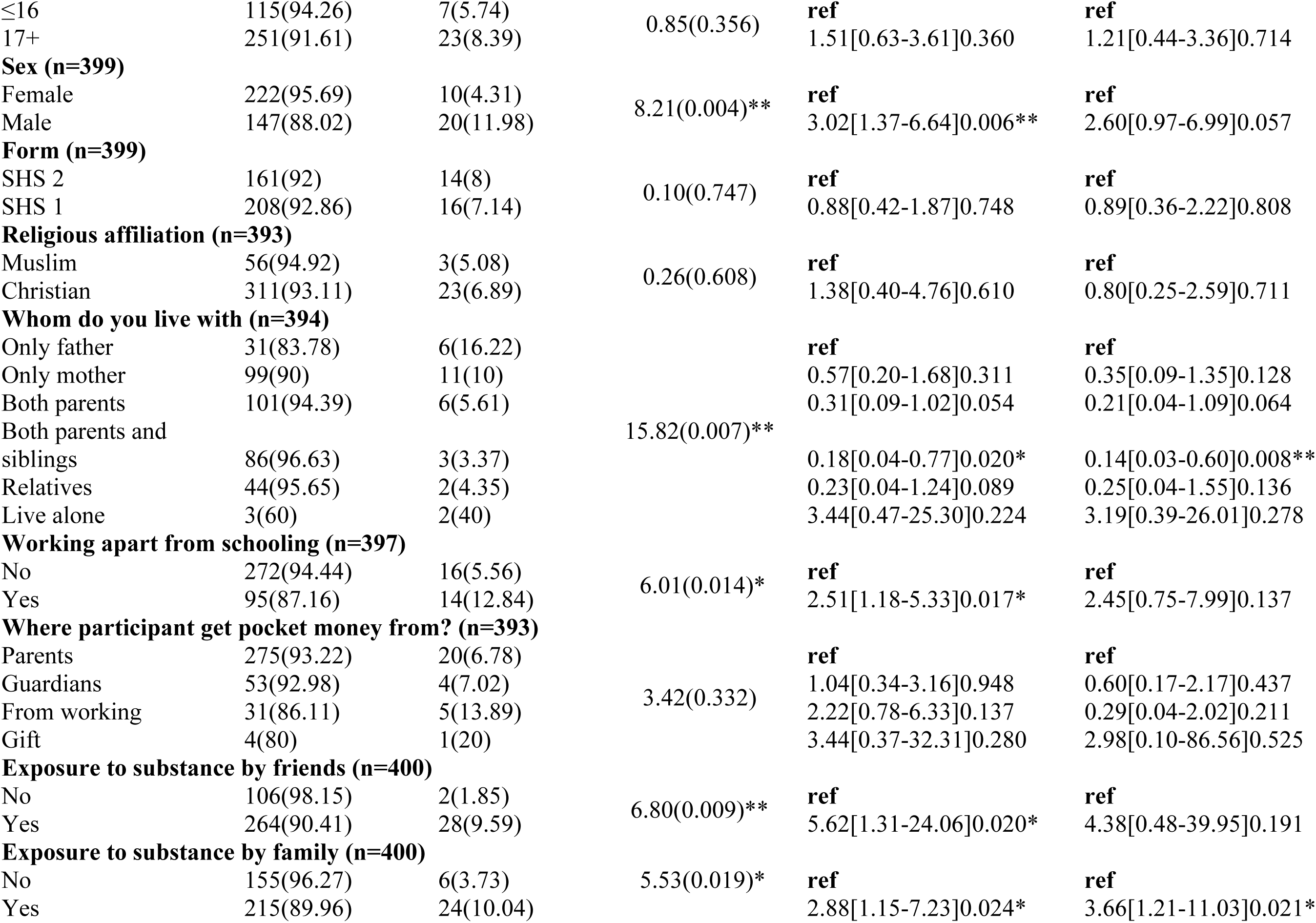
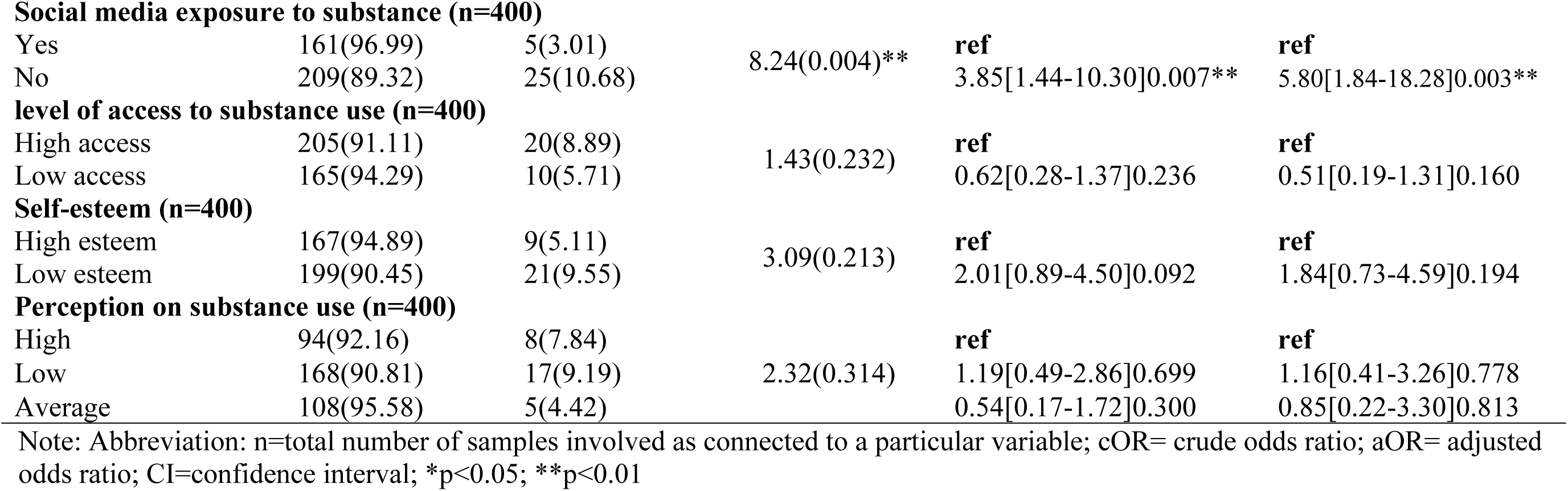
Personal characteristics, social and intrapersonal factors, and marijuana use.

### Substance use dependence

Substance use dependence was assessed using a scale ranging from 0 (no dependence) to 4 (high dependence), with a score of two or higher indicating significant dependence. As shown in Table 7, a substantial proportion of alcohol users (62.03%, n=49) met the criteria for alcohol dependence, while the remaining 37.97% (n=30) did not. Among marijuana users, 66.67% (n=20) demonstrated dependence on the substance. Shisha dependence was notably high, with 80.36% (n=41) of shisha users meeting the criteria for dependence. In contrast, cigarette dependence was observed in 46.67% of cigarette users.

**Table 7.** Substance use dependence.

| Variable | Type of substance use dependence |
| --- | --- |

|  | Alcohol |  |  | Cigarette |  |  | Shisha |  |  | Marijuana |  |  |
| --- | --- | --- | --- | --- | --- | --- | --- | --- | --- | --- | --- | --- |
| | Yes (2-4) | No ($\leq 1$) | Total | Yes (2-4) | No ($\leq 1$) | Total | Yes (2-4) | No ($\leq 1$) | Total | Yes (2-4) | No ($\leq 1$) | Total |
|  | n(%) | n(%) |  | n(%) | n(%) |  | n(%) | n(%) |  | n(%) | n(%) |  |
| Overall | 49(62.03) | 30(37.97) | 79 | 7(46.67) | 8(53.33) | 15 | 41(80.36) | 10(19.61) | 51 | 20(66.67) | 10(33.33) | 30 |
| **Age group** |  |  |  |  |  |  |  |  |  |  |  |  |
| $\leq 16$ | 13(59.09) | 9(40.91) | 22 | 2(50.00) | 2(50.00) | 4 | 7(87.50) | 1(12.50) | 8 | 4(57.14) | 3(42.86) | 7 |
| 17+ | 36(63.16) | 21(36.84) | 57 | 5(45.45) | 6(54.55) | 11 | 34(79.07) | 9(20.93) | 43 | 16(69.57) | 7(30.43) | 23 |
| Total | 49(62.03) | 30(37.97) | 79 | 7(46.67) | 8(53.33) | 15 | 41(80.39) | 10(19.61) | 51 | 20(66.67) | 10(33.33) | 30 |
| **Sex** |  |  |  |  |  |  |  |  |  |  |  |  |
| Male | 28(62.22) | 17(37.78) | 45 | 4(50.00) | 4(50.00) | 8 | 24(88.89) | 3(11.11) | 27 | 14(70.00) | 6(30.00) | 20 |
| Female | 21(61.76) | 13(38.24) | 34 | 3(42.86) | 4(57.14) | 7 | 17(70.83) | 7(29.17) | 24 | 6(60.00) | 4(40.00) | 10 |
| Total | 49(62.03) | 30(37.97) | 79 | 7(46.67) | 8(53.33) | 15 | 41(80.39) | 10(19.61) | 51 | 20(66.67) | 10(33.33) | 30 |
| **Form** |  |  |  |  |  |  |  |  |  |  |  |  |
| SHS 1 | 35(64.81) | 19(35.19) | 54 | 3(33.33) | 6(66.67) | 9 | 26(81.25) | 6(18.75) | 32 | 12(75.00) | 4(25.00) | 16 |
| SHS 2 | 14(58.33) | 10(41.67) | 24 | 4(66.67) | 2(33.33) | 6 | 15(78.95) | 4(21.05) | 19 | 8(57.14) | 6(42.86) | 14 |
| Total | 49(62.82) | 29(37.18) | 78 | 7(46.67) | 8(53.33) | 15 | 41(80.39) | 10(19.61) | 51 | 20(66.67) | 10(33.33) | 30 |
| **Religious affiliation** |  |  |  |  |  |  |  |  |  |  |  |  |
| Christian | 45(63.38) | 26(36.62) | 71 | 5(41.67) | 7(58.33) | 12 | 35(81.40) | 8(18.60) | 43 | 14(60.87) | 9(39.13) | 23 |
| Muslim | 3(42.86) | 4(57.14) | 7 | 1(50.00) | 1(50.00) | 2 | 4(66.67) | 2(33.33) | 6 | 3(100.00) | 0(0.00) | 3 |
| Total | 48(61.54) | 30(38.46) | 78 | 6(42.86) | 8(57.14) | 14 | 39(79.59) | 10(20.41) | 49 | 17(65.38) | 9(34.62) | 26 |

## Discussion

This study examined substance use patterns and their correlates among Ghanaian in-school adolescents. We found that cigarettes, marijuana, shisha, and alcohol were the primary substances used by participants. This aligns with findings from Chile, where cigarettes, alcohol, and cannabis were common among adolescents (33). However, alcohol was the most prevalent substance in our study, contrasting with an Iranian review that identified cigarettes and hookah (shisha) as the most common (34). Notably, our alcohol prevalence was lower than the 34.63% observed among adolescents in 47 European countries (35), potentially due to their larger, cross-national sample compared to our two-school study and regional differences between settings.

Marijuana prevalence in our study (7.5%) exceeded the 3.4% found in a Ghanaian study using 2012 Global School-Based Student Health Survey data (14,36), but was lower than the 9% reported among junior high school students (37). These variations may stem from differences in sample size and the number of schools involved.

Shisha use prevalence (12.59%) was higher than the 10.3% observed in Ghanaian urban slums and communities (20), yet lower than the 13.4% reported among Sudanese youth (38). These discrepancies likely reflect the distinct study settings and populations. This high prevalence is concerning, given the potential negative consequences of substance use on these students and the association of high school shisha use with deviant behaviours (39).

Our study’s cigarette use prevalence (3.77%) was lower than the 11.1% reported in Nigeria (40). This might be attributed to setting differences and the global decline in tobacco consumption (12). Friends, online/social media, markets, and drug peddlers were identified as sources of these substances. The influence of peers and the lack of enforcement of existing policies and regulations likely contribute to the availability of these substances among high school students (20).

Our study confirmed the association between various personal characteristics, social, and intrapersonal factors, and substance use. Alcohol use was more prevalent among males, aligning with previous studies in Morocco and Ethiopia (41,42). Further, living with relatives and having friends who consume alcohol were significant predictors, emphasizing the importance of peer influence and social networks in adolescent substance use (Bondah et al., 2020; Henneberger et al., 2021; Osaki et al., 2018; Quiroga et al., 2018; Reda et al., 2012).

While sex was not significantly associated with cigarette use in our study, it was in a Nigerian study (40). Although peer use is often a reliable indicator, we found no association, contrasting a study in Ethiopia that found a significant association with having smoker friends and parents (47). However, our finding of a significant association with family exposure to cigarette use aligns with their results.

Age was associated with shisha smoking, consistent with a Lebanese study (48), but not with studies in Sudan and the UK (38,49). We found more boys smoked shisha, though not significantly, contrasting a Lebanese study finding more girls (50). This discrepancy could be due to differing social contexts and gendered expectations (51).

Our study aligns with (52) in finding family and friend shisha use as predictors. While initially significant, low perception of consequences became insignificant after controlling for other factors, contrasting a Canadian study where low perception increased the odds of shisha use (53). This could be due to misconceptions about shisha’s harmfulness (8).

Finally, our study revealed significant associations between marijuana use and family exposure, non-exposure to social media, and living with both parents and siblings, echoing the importance of family influence found in Moroccan and Northern Irish studies (54,55). While initially significant, being male, peer exposure, and working alongside schooling became insignificant after adjustment, contrasting studies linking these factors to marijuana use (18,37). This could be due to our study’s smaller sample size compared to larger studies finding age, low self-esteem, and substance availability as significant predictors (6,56).

The current study revealed a notable prevalence of substance dependence among students who reported using various substances. Specifically, we observed high rates of dependence on alcohol (62.03%), shisha (80.36%), and marijuana (66.67%), while dependence on cigarettes was comparatively lower (46.67%). This lower rate of cigarette dependence aligns with reports of decreased cigarette smoking in Ghana (57).

Our findings echo previous research indicating substance dependence among adolescents and young adults. Similar patterns of marijuana and alcohol dependence in 12th-grade students have been found in a previous study (58). Furthermore, studies conducted in diverse geographical contexts, such as Egypt (59,60) Jordan (61), and Romania (62), have consistently demonstrated substance dependence among student populations, particularly with regard to tobacco.

Taken together, these findings underscore the global nature of substance dependence among students and highlight the need for targeted interventions and prevention strategies aimed at reducing substance use and associated harms in this vulnerable population.

### Limitations of the study

This study has several limitations. The exclusion of Senior High School 3 (SHS 3) students may underestimate the prevalence and associated factors of substance use. Reliance on self-reporting could lead to underreporting due to social desirability and recall bias. The cross-sectional design also prevents establishing causality. Future research should include a wider age range, use a longitudinal design, and incorporate objective measures of substance use to address these limitations.

## Conclusion

This study found that substance use is prevalent among senior high school students in the Accra Metropolitan Area. Cigarettes, marijuana, shisha, and alcohol are the most commonly used substances, with varying prevalence rates. The primary source of these substances is friends, suggesting that students are bringing them to school without the knowledge of authorities.

Several socio-demographic factors, including substance use among friends and family, as well as sex, are associated with substance use among students. Notably, males are more likely to use substances compared to females.

Furthermore, a significant proportion of students who use substances exhibit signs of dependence. This is particularly evident in SHS 1 students for most substances, except cigarettes. Additionally, males and students older than 17 years are more likely to be dependent on these substances.

These findings underscore the need for targeted interventions and prevention programs to address substance use and dependence among senior high school students in the Accra Metropolitan Area. Further research is needed to explore the underlying reasons for substance use, the impact of dependence, and the effectiveness of various prevention and intervention strategies.

## Data Availability

If the data are all contained within the manuscript and/or Supporting Information files, enter the following: All relevant data are within the manuscript and its Supporting Information files.

## Acknowledgments

Our sincere appreciation goes to the school authorities who permitted this study to be conducted in their schools and the students who participated in the study. Without them this study would not have been possible.

